# Effectiveness of Vaccination and Previous Infection Against Omicron Infection and Severe Outcomes in Children Under 12 Years of Age

**DOI:** 10.1101/2023.01.18.23284739

**Authors:** Dan-Yu Lin, Yangjianchen Xu, Yu Gu, Donglin Zeng, Bradford Wheeler, Hayley Young, Zack Moore, Shadia K Sunny

**Author notes:** Address correspondence to Dr. Dan-Yu Lin, Department of Biostatistics, University of North Carolina, Chapel Hill, NC 27599-7420, USA.

## Abstract

**Background:** Data on the protection conferred by Covid-19 vaccination and previous SARS-CoV-2 infection against omicron infection and severe outcomes in children can inform prevention strategies.

**Methods:** We obtained vaccination records and clinical outcomes for 1,368,721 North Carolina residents 11 years of age or younger from October 29, 2021 to January 6, 2023. We used Cox regression to estimate the time-varying effects of primary and booster vaccination and previous infection on the risks of omicron infection, hospitalization, and death.

**Results:** For children 5–11 years of age, the effectiveness of primary vaccination against infection was 59.9% (95% confidence interval [CI], 58.5 to 61.2), 33.7% (95% CI, 32.6 to 34.8), and 14.9% (95% CI, 12.3 to 17.5) at 1, 4 and 10 months after the first dose; the effectiveness of a monovalent or bivalent booster dose after 1 month was 24.4% (95% CI, 14.4 to 33.2) or 76.7% (95% CI, 45.7 to 90.0); and the effectiveness of omicron infection against reinfection was 79.9% (95% CI, 78.8 to 80.9) and 53.9% (95% CI, 52.3 to 55.5) after 3 and 6 months, respectively. For children 0–4 years of age, the effectiveness of primary vaccination against infection was 63.8% (95% CI, 57.0 to 69.5) and 58.1% (95% CI, 48.3 to 66.1) at 2 and 5 months after the first dose, and the effectiveness of omicron infection against reinfection was 77.3% (95% CI, 75.9 to 78.6) and 64.7% (95% CI, 63.3 to 66.1) after 3 and 6 months, respectively. For both age groups, vaccination and previous infection had better effectiveness against hospitalization and death than against infection.

**Conclusions:** Covid-19 vaccination was effective against omicron infection and severe outcomes in children under the age of 12 years, although the effectiveness decreased over time. Bivalent boosters were more effective than monovalent boosters. Immunity acquired via omicron infection was very high and waned gradually over time.

---

Omicron is highly transmissible and can cause severe illness, especially in immunocompromised individuals. Covid-19 vaccination and previous SARS-CoV-2 infection have been shown to reduce the risk of omicron infection and severe outcomes in adults and adolescents.^1-13^ However, the data on the effectiveness of Covid-19 vaccination in children under 12 years of age are limited, and the effects of previous SARS-CoV-2 infection on omicron infection in these children are not well understood.^3-4,12,14-17^ Using surveillance data on Covid-19 vaccination and disease incidence from the state of North Carolina as of June 3, 2022, we estimated the effects of the BNT162b2 (Pfizer–BioNTech) vaccine and previous SARS-CoV-2 infection on infection with omicron’s BA.1 and BA.2 lineages among children 5–11 years old.^18^ Due to limited follow-up, the long-term effects of vaccination and previous infection were unclear.

This article presents the North Carolina surveillance data on Covid-19 vaccination and clinical outcomes in children younger than 12 years over the entire omicron period, up until January 6, 2023. The expanded dataset allowed characterization of the long-term effects of primary vaccination and previous SARS-CoV-2 infection on omicron infection and severe outcomes among children 5–11 years of age. It also allowed comparison of the effectiveness of monovalent and bivalent boosters in this age group. Finally, it allowed estimation of the time-varying effects of primary vaccination and previous SARS-CoV-2 infection on omicron infection and severe outcomes among children younger than 5 years. This study covered all lineages of the omicron variant, including BA.1, BA.2, BA.4, BA.5, BQ.1/BQ.1.1, and XBB/XBB.1.5.

## Methods

### Data Sources

#### North Carolina COVID-19 Surveillance System

The North Carolina Covid-19 Surveillance System (NC COVID) is a web-based central repository of person-level laboratory and communicable-disease investigation data. Covid-19 cases are populated according to lab reports from clinical laboratories that are mandated to report results. At-home testing results are not included. The database contains positive SARS-CoV-2 test results for all cases and index reinfections using the unique person identifier and person-event infection variables. Covid-19-related hospitalizations and deaths are documented through local health department case investigation. For cases reported on January 1 of 2022 and going forward, vital record criteria have been used to expand the Covid-19 death surveillance.

#### COVID-19 Vaccine Management System

The Covid-19 Vaccine Management System (CVMS) is a secure, cloud-based system that tracks information about provider enrollment and vaccine products administered, schedules appointments according to the recommended vaccination schedule, and allows the state to manage vaccine supply. The CVMS records are transferred daily to the North Carolina Department of Health and Human Services Business Intelligence Data Platform, where the data are processed to create a recipient-based view of Covid-19 vaccination history.

### Study Design

#### We considered children in two age groups

5–11 years old and 0–4 years old. The US Food and Drug Administration (FDA) authorized the BNT162b2 vaccine for emergency use in children 5 through 11 years of age on October 29, 2021 and expanded eligibility for the BNT162b2 vaccine booster dose to this age group on May 17, 2022.^19-20^ In addition, the FDA authorized the mRNA-1273 (Moderna) and BNT162b2 vaccines for children as young as 6 months of age on June 17, 2022.^21^

We extracted individual-level data on vaccination histories from the authorization date of October 29, 2021 to the study end date of January 6, 2023 for children 5–11 years of age and from the authorization date of June 17, 2022 to January 6, 2023 for children 0–4 years of age, as well as individual-level data on SARS-CoV-2 infection, hospitalization, and death from March 11, 2020 to January 6, 2023 for children 0 through 11 years of age, by linking the NC COVID and CVMS databases through a Master Patient Index. Of note, infection was both an exposure and an outcome, in that we were studying the effects of previous infection on future infection and severe outcomes. Finally, we used the 2020 Bridged-Race Population estimates produced by the US Census Bureau to determine the total number of residents with each combination of demographic variables, i.e., age, sex, race/ethnicity, geographic region (Coastal Plain, Piedmont, Mountains), and county-level vaccination rate.

### Statistical Analysis

#### We considered two types of outcomes

SARS-CoV-2 infection and composite endpoint of hospitalization and death. The latter was defined as time to severe SARS-CoV-2 infection that results in hospitalization or death. We treated SARS-CoV-2 infection as a recurrent event and related the rate of infection to immunity-conferring events (i.e., vaccination and previous infection) through the proportional rates model.^22^ In addition, we related the hazard of severe SARS-CoV-2 infection to immunity-conferring events through the proportional hazards model.^23^

The effect of each exposure (i.e., vaccination or previous infection) on the risk of SARS-CoV-2 infection or severe SARS-CoV-2 infection was characterized by a time-varying rate ratio or hazard ratio, respectively, as a function of the time elapsed since the exposure; stratification by the date of exposure and interactions between the two exposures were allowed.^8,18^ We measured each event time from the start of the study to control for time-varying confounders (e.g., circulating strains, use of masks, school opening) by comparing disease incidence between vaccinated and unvaccinated children, as well as between previously infected and uninfected children, on the same date.^8,18^ We included demographic variables (age, sex, race/ethnicity, geographic region, county-level vaccination rate) as covariates to adjust for potential confounding by individual characteristics and geographic location.

In the first set of analyses, we estimated the effects of primary vaccination and previous SARS-CoV-2 infection on the two types of outcomes in children 5–11 years of age. We estimated the overall effects of these two exposures, as well as the effects of vaccination on infection by date of first dose and by previous infection status and the effects of previous infection on future infection by virus strain and by vaccination status.

In the second set of analyses, we estimated the effectiveness of booster vaccination and compared monovalent and bivalent boosters in children 5–11 years of age. We included the date of primary series as an additional covariate.

The third set of analyses was similar to the first, but for children 0–4 years of age. We also compared the mRNA-1273 and BNT162b2 vaccines.

The effectiveness of vaccination and previous infection was defined by one minus the rate ratio or hazard ratio, multiplied by 100%. The parameters in each model were estimated by maximizing the partial likelihood with potentially censored observations.^8^ Corresponding 95% confidence intervals were constructed.

## Results

### Study Population

Table 1 summarizes the demographic characteristics, vaccine uptakes, and clinical outcomes of the study participants. As of January 6, 2023, a total of 39,261, 216,330, and 46,895 children 5– 11 years of age had received only one dose, only two doses, and three doses of an mRNA vaccine, respectively; and a total of 11,235, 28,066, and 11,529 children 0–4 years of age had received only one dose, only two doses, and three doses of an mRNA vaccine, respectively. For the latter age group, the third dose was part of the primary series for immune-compromised children, rather than as a booster dose.

**Table 1.**
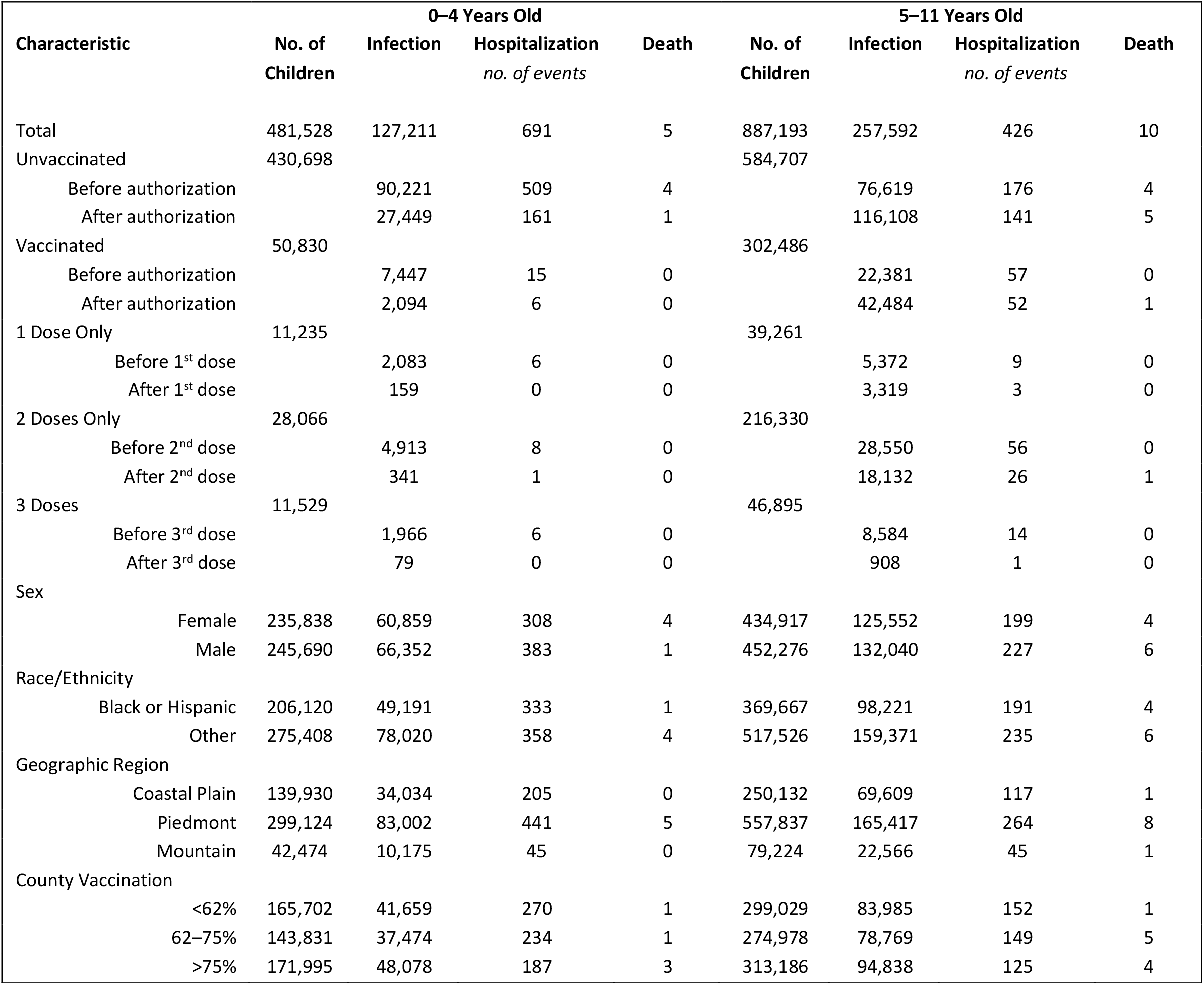
Demographic and Clinical Characteristics of Study Participants.

| Characteristic | No. of Children | 0–4 Years Old |  |  | No. of Children | 5–11 Years Old |  |  |
| --- | --- | --- | --- | --- | --- | --- | --- | --- |
|  |  | Infection | Hospitalization<br>*no. of events* | Death |  | Infection | Hospitalization<br>*no. of events* | Death |
| Total | 481,528 | 127,211 | 691 | 5 | 887,193 | 257,592 | 426 | 10 |
| Unvaccinated | 430,698 |  |  |  | 584,707 |  |  |  |
| Before authorization |  | 90,221 | 509 | 4 |  | 76,619 | 176 | 4 |
| After authorization |  | 27,449 | 161 | 1 |  | 116,108 | 141 | 5 |
| Vaccinated | 50,830 |  |  |  | 302,486 |  |  |  |
| Before authorization |  | 7,447 | 15 | 0 |  | 22,381 | 57 | 0 |
| After authorization |  | 2,094 | 6 | 0 |  | 42,484 | 52 | 1 |
| 1 Dose Only | 11,235 |  |  |  | 39,261 |  |  |  |
| Before 1<sup>st</sup> dose |  | 2,083 | 6 | 0 |  | 5,372 | 9 | 0 |
| After 1<sup>st</sup> dose |  | 159 | 0 | 0 |  | 3,319 | 3 | 0 |
| 2 Doses Only | 28,066 |  |  |  | 216,330 |  |  |  |
| Before 2<sup>nd</sup> dose |  | 4,913 | 8 | 0 |  | 28,550 | 56 | 0 |
| After 2<sup>nd</sup> dose |  | 341 | 1 | 0 |  | 18,132 | 26 | 1 |
| 3 Doses | 11,529 |  |  |  | 46,895 |  |  |  |
| Before 3<sup>rd</sup> dose |  | 1,966 | 6 | 0 |  | 8,584 | 14 | 0 |
| After 3<sup>rd</sup> dose |  | 79 | 0 | 0 |  | 908 | 1 | 0 |
| Sex |  |  |  |  |  |  |  |  |
| Female | 235,838 | 60,859 | 308 | 4 | 434,917 | 125,552 | 199 | 4 |
| Male | 245,690 | 66,352 | 383 | 1 | 452,276 | 132,040 | 227 | 6 |
| Race/Ethnicity |  |  |  |  |  |  |  |  |
| Black or Hispanic | 206,120 | 49,191 | 333 | 1 | 369,667 | 98,221 | 191 | 4 |
| Other | 275,408 | 78,020 | 358 | 4 | 517,526 | 159,371 | 235 | 6 |
| Geographic Region |  |  |  |  |  |  |  |  |
| Coastal Plain | 139,930 | 34,034 | 205 | 0 | 250,132 | 69,609 | 117 | 1 |
| Piedmont | 299,124 | 83,002 | 441 | 5 | 557,837 | 165,417 | 264 | 8 |
| Mountain | 42,474 | 10,175 | 45 | 0 | 79,224 | 22,566 | 45 | 1 |
| County Vaccination |  |  |  |  |  |  |  |  |
| <62% | 165,702 | 41,659 | 270 | 1 | 299,029 | 83,985 | 152 | 1 |
| 62–75% | 143,831 | 37,474 | 234 | 1 | 274,978 | 78,769 | 149 | 5 |
| >75% | 171,995 | 48,078 | 187 | 3 | 313,186 | 94,838 | 125 | 4 |

Between the vaccination authorization date of October 29, 2021 and the study end date of January 6, 2023, the 584,707 unvaccinated children 5–11 years of age experienced 116,108 SARS-CoV-2 infections, 141 of which were known to result in hospitalization and 5 of which were known to result in death, whereas the 302,486 vaccinated children 5–11 years of age experienced 42,484 SARS-CoV-2 infections, 52 of which were known to result in hospitalization and only 1 was known to result in death. Between the vaccine authorization date of June 17, 2022 and January 6, 2023, the 430,698 unvaccinated children 0–4 years of age experienced 27,449 SARS-CoV-2 infections, 161 of which were known to result in hospitalization and 1 was known to result in death, whereas the 50,830 vaccinated children 0–4 years of age experienced 2,094 SARS-CoV-2 infections, 6 of which were known result in hospitalization and none of which were known to result in death.

### Primary Vaccination in Children 5–11 Years of Age

Figure 1 shows the effectiveness of the primary vaccine series over time in children 5–11 years of age. The effectiveness against infection reached a peak level of 59.9% (95% CI, 58.5 to 61.2) at 1 month after the first dose, and it decreased to 33.7% (95% CI, 32.6 to 34.8) at 4 months and to 14.9% (95% CI, 12.3 to 17.5) at 10 months (Fig. 1A, Table S1). The ramping and waning patterns were broadly similar among children who were vaccinated on different dates, and the emergence of the BQ.1/BQ.1.1 and XBB/XBB.1.5 strains seemed to reduce vaccine effectiveness (Fig. 1B, Table S2).

**Figure 1.**
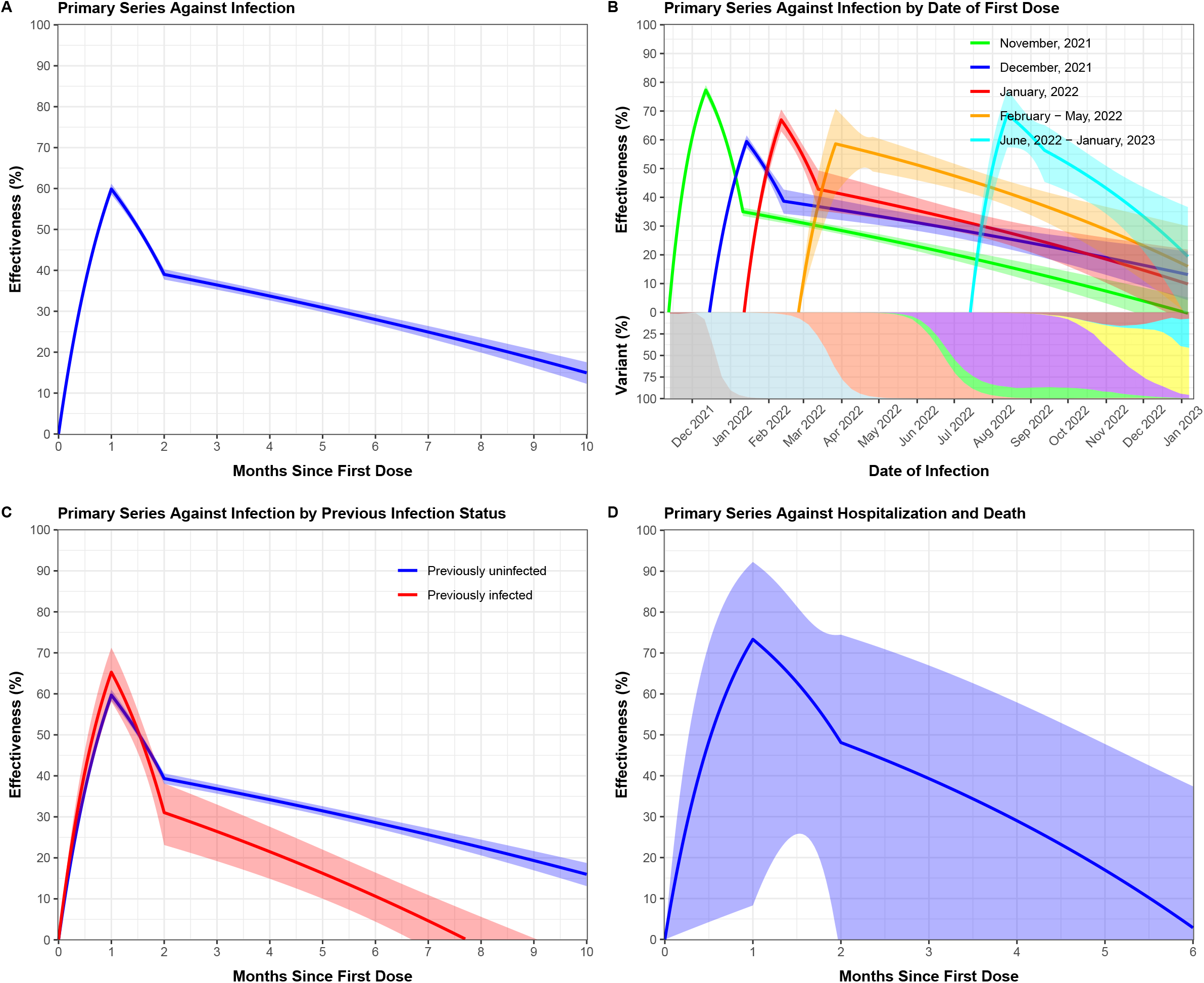
Effectiveness of two doses of an mRNA vaccine, compared with no vaccination, against omicron infection and severe outcomes in children 5 to 11 years of age: (A) vaccine effectiveness against infection; (B) vaccine effectiveness against infection by date of first dose; (C) vaccine effectiveness against infection by previous infection status; and (D) vaccine effectiveness against hospitalization and death. In (B), each curve starts at the median date of the first dose for the children in that cohort, and the proportions of the delta variant and the omicron variant’s BA.1, BA.2, BA.4, BA.5, BQ.1/BQ.1.1, XBB/XBB.1.5, and other strains are indicated by the gray, light blue, coral, green, purple, yellow, pink, and brown areas, respectively. In (C), effectiveness is calculated for vaccination alone given the status of previous infection. Estimates of effectiveness are shown by solid curves, and 95% confidence intervals are shown by shaded bands.

Among previously infected children, the effectiveness of primary vaccination against infection reached a peak level of 65.3% (95% CI, 58.1 to 71.2) at 1 month and declined to 26.4% (95% CI, 19.2 to 33.0) at 3 months and to 10.6% (95% CI, 4.4 to 16.4) at 6 months; among previously uninfected children, the effectiveness reached a peak level of 59.7% (95% CI, 58.3 to 61.0) at 1 month and declined to 36.8% (95% CI, 35.7 to 38.0) at 3 months and to 28.6% (95% CI, 27.3 to 29.9) at 6 months (Fig. 1C, Table S3).

The effectiveness of primary vaccination against hospitalization and death reached a peak level of 73.3% (95% CI, 8.3 to 92.3) at 1 month and waned afterward (Fig. 1D, Table S4). The estimates were highly variable due to the small number of events.

### Previous Infection in Children 5–11 Years of Age

Figure 2 shows the effectiveness of previous SARS-CoV-2 infection over time in children 5–11 years of age. The effectiveness of previous infection with the omicron variant against reinfection with the omicron variant was 79.9% (95% CI, 78.8 to 80.9) at 3 months and 53.9% (95% CI, 52.3 to 55.5) at 6 months (Fig. 2A, Table S5). The long-term effectiveness was higher among vaccinated children than among unvaccinated children (Fig. 2B, Table S6). The effectiveness of previous infection with the delta variant against future infection was similar between vaccinated and unvaccinated children (Fig. 2C, Table S7). The effectiveness of previous infection against severe reinfection resulting in hospitalization and death was 83.8% (95% CI, 59.1 to 93.6) at 3 months, 76.2% (95% CI, 49.4 to 88.8) at 6 months, and 64.9% (95% CI, 34.9 to 81.1) at 9 months (Fig. 2D, Table S8).

**Figure 2.**
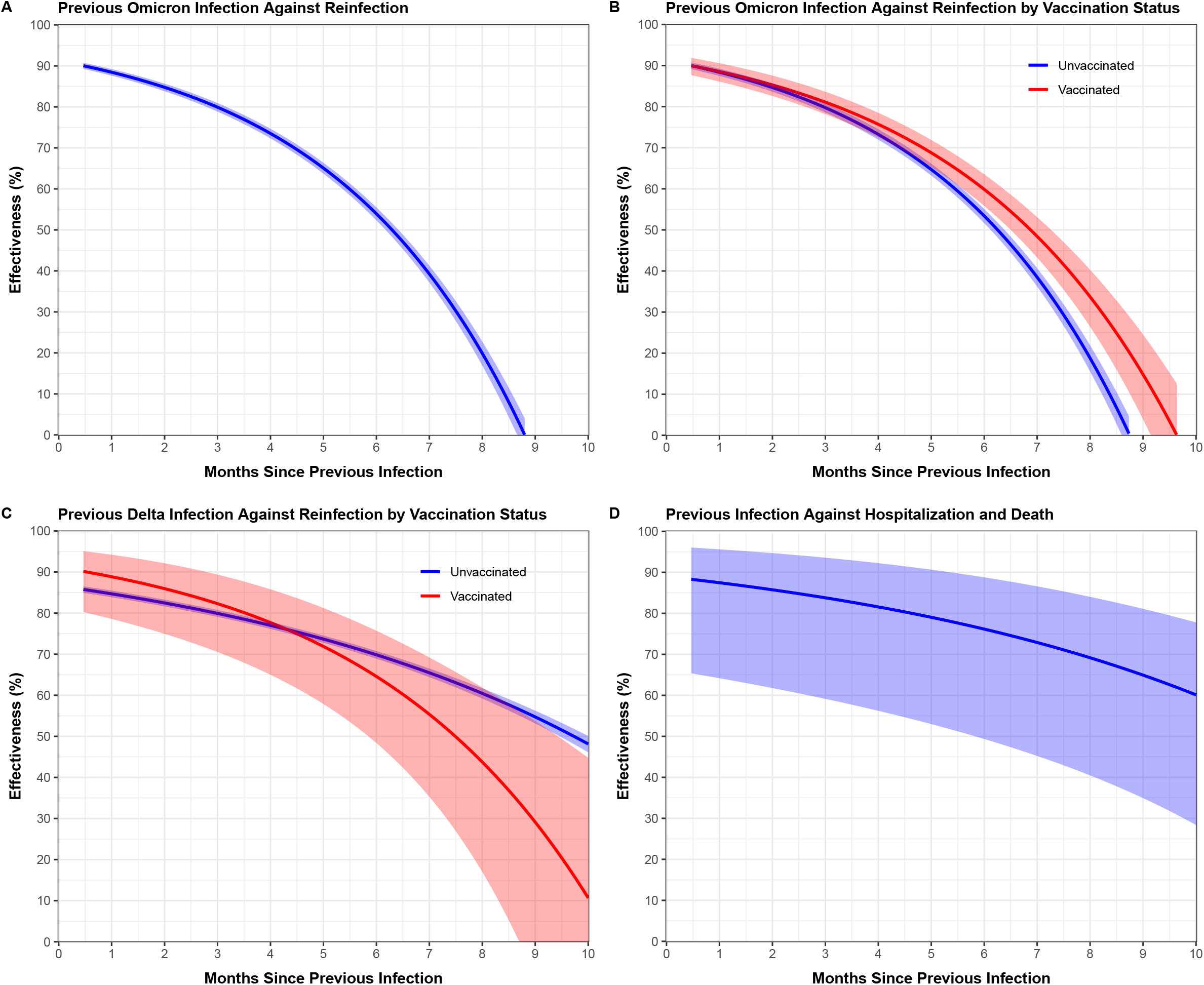
**Effectiveness of previous SARS-CoV-2 infection, compared with no previous infection, against future infection and severe outcomes in children 5 to 11 years of age: (A) effectiveness of previous omicron infection against omicron reinfection; (B) effectiveness of previous omicron infection against omicron reinfection by vaccination status; (C) effectiveness of previous infection with the delta variant against reinfection by vaccination status; and (D) effectiveness of previous infection against severe reinfection resulting in hospitalization or death.** In (B) and (C), effectiveness is calculated for previous infection alone given the vaccination status. Estimates of effectiveness are shown by solid curves, and 95% confidence intervals are shown by shaded bands.

### Booster Dose in Children 5–11 Years of Age

Figure 3 shows the effectiveness of a booster dose, compared with two doses, against SARS-CoV-2 infection in children 5–11 years of age. The effectiveness of a monovalent booster dose was 24.4% (95% CI, 14.4 to 33.2) after 1 month and 23.1% (95% CI, 15.6 to 30.0) after 2 months, whereas that of a bivalent booster dose was 76.7% (95% CI, 45.7 to 90.0) after 1 month and 47.3% (95% CI, -17.9 to 76.4) after 2 months (Table S9). We were unable to obtain an accurate estimate for the booster effectiveness against hospitalization and death because there was only 1 hospitalization and no death after receipt of a booster dose (Table 1).

**Figure 3.**
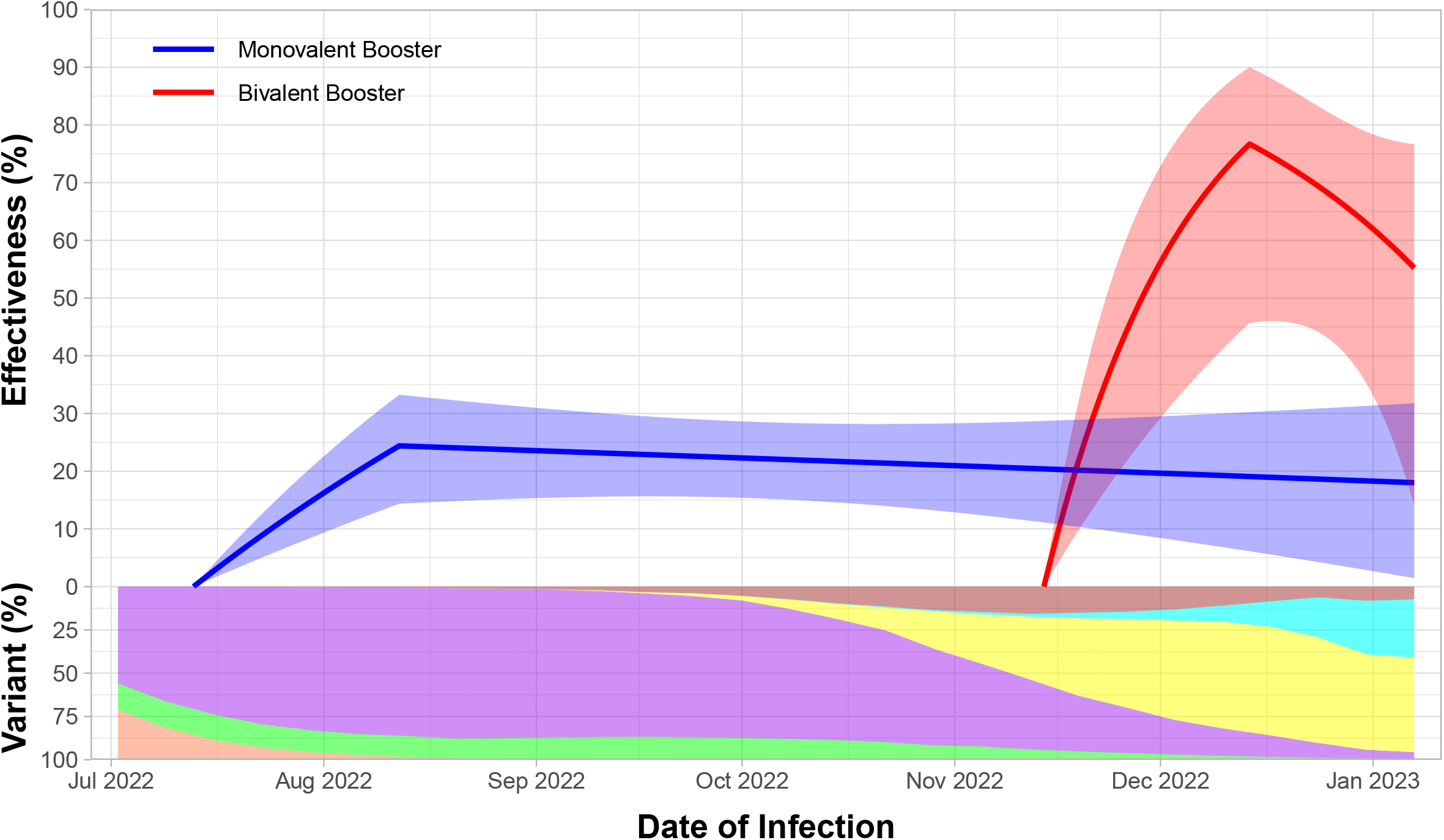
Effectiveness of a monovalent or bivalent booster dose, compared with two monovalent doses, against omicron infection in children 5 to 11 years of age. The proportions of BA.2, BA.4, BA.5, BQ.1/BQ.1.1, and XBB/XBB.1.5, and other strains are indicated by the coral, green, purple, yellow, pink, and brown areas, respectively. Estimates of effectiveness are shown by solid curves, and 95% confidence intervals are shown by shaded bands.

### Primary Vaccination in Children 0–4 Years of Age

Figure 4 (A-D) shows the effectiveness of a primary vaccine series against SARS-CoV-2 infection over time in children 0–4 years of age. The effectiveness reached a peak level of 63.8% (95% CI, 57.0 to 69.5) at 2 months after the first dose and decreased to 58.1% (95% CI, 48.3 to 66.1) at 5 months (Fig. 4A, Table S10). The ramping and waning patterns were broadly similar among children who were vaccinated on different dates (Fig. 4B, Table S11). Both the mRNA-1273 and BNT162b2 vaccines were effective (Fig. 4C, Table S12). Vaccination was effective for both previously infected and previously uninfected children (Fig. 4D, Table S13).

**Figure 4.**
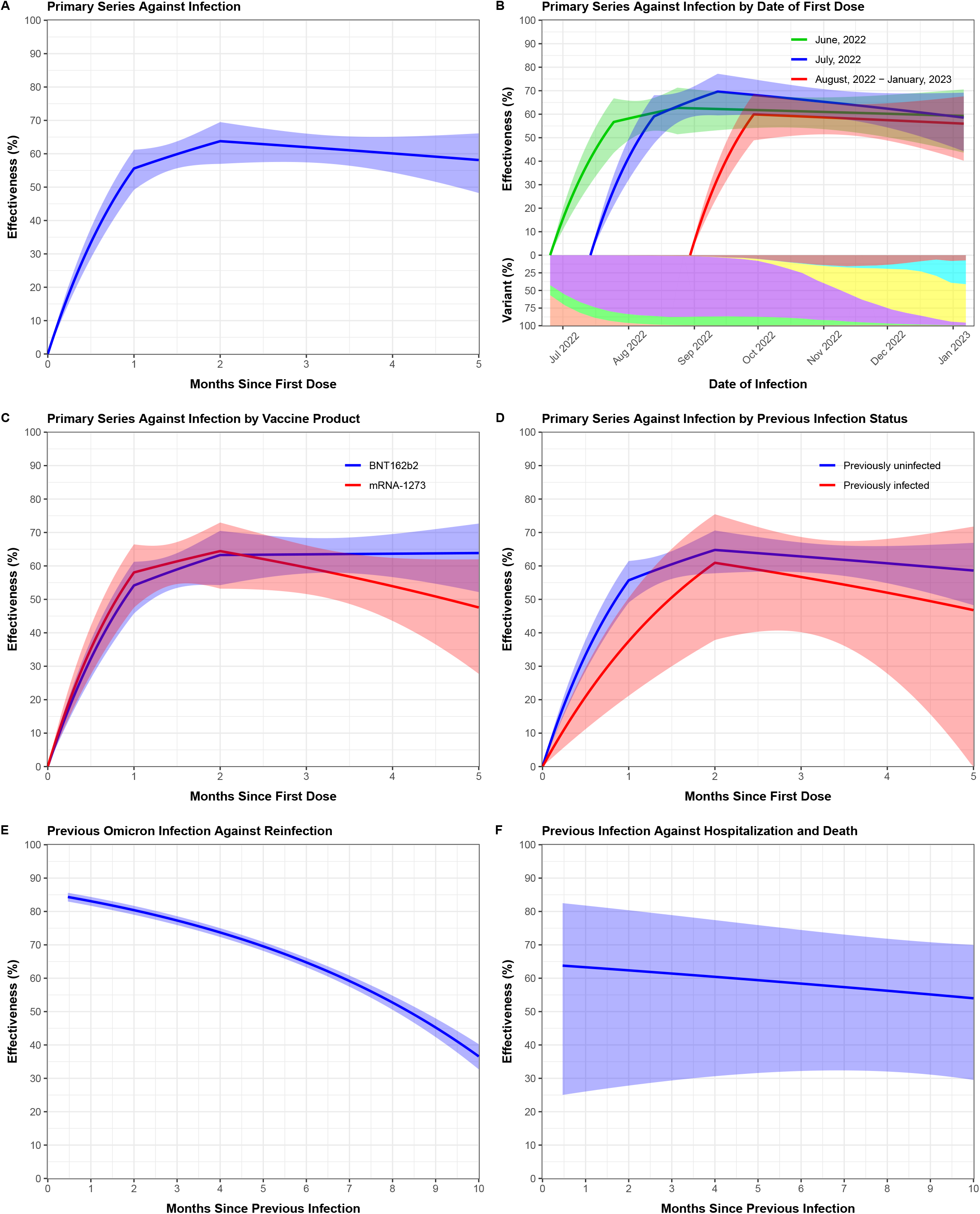
**Effectiveness of two doses of an mRNA vaccine, compared with no vaccination, and effectiveness of previous SARS-CoV-2 infection, compared with no previous infection, against subsequent infection and severe outcomes in children 0 to 4 years of age: (A) vaccine effectiveness against infection; (B) vaccine effectiveness against infection by date of first dose; (C) vaccine effectiveness against infection by vaccine product; (D) vaccine effectiveness against infection by previous infection status; (E) effectiveness of omicron infection against reinfection; and (F) effectiveness of previous infection against severe reinfection resulting in hospitalization or death.** In (B), each curve starts at the median date of the first dose for the children in that cohort, and the proportions of BA.2, BA.4, BA.5, BQ.1/BQ.1.1, and XBB/XBB.1.5, and other strains are indicated by the coral, green, purple, yellow, pink, and brown areas, respectively. In (D), effectiveness is calculated for vaccination alone given the status of previous infection. Estimates of effectiveness are shown by solid curves, and 95% confidence intervals are shown by shaded bands.

Only one of the 42,983 vaccinated children was known to be hospitalized after vaccination and none were known to die (Table 1).

### Previous Infection in Children 0–4 Years of Age

Figure 4 (E-F) shows the effectiveness of previous SARS-CoV-2 infection over time in children 0– 4 years of age. The effectiveness of previous infection with the omicron variant against reinfection with the omicron variant was 77.3% (95% CI, 75.9 to 78.6), 64.7% (95% CI, 63.3 to 66.1), and 45.2% (95% CI, 42.4 to 48.0) at 3 months, 6 months, and 9 months, respectively (Fig. 4E, Table S14). The effectiveness of previous infection against severe reinfection resulting in hospitalization or death was 61.4% (95% CI, 29.4 to 78.9), 58.4% (95% CI, 32.2 to 74.5), and 55.1% (95% CI, 31.1 to 70.8) at 3 months, 6 months, and 9 months, respectively (Fig. 4F, Table S15).

## Discussion

This study yielded important findings about Covid-19 vaccination and previous SARS-CoV-2 infection in children under 12 years of age. First, vaccination was effective against omicron infection, hospitalization, and death, although the effectiveness waned over time. Second, both the mRNA-1273 and BNT162b2 vaccines were effective. Third, bivalent boosters were more effective than monovalent boosters. Fourth, previous SARS-CoV-2 infection induced strong immunity against future infection, although the immunity waned gradually over time. Fifth, vaccination provided additional protection for previously infected children, and omicron infection induced strong immunity in both vaccinated and unvaccinated children. Finally, the effectiveness of both vaccination and previous infection was higher for children under 5 years of age than for children 5–11 years of age.

Our estimates for the effectiveness of vaccination in children under 12 years of age were lower than those of adults and adolescents previously reported.^1-12,24-26^ However, vaccines were administered in children under the age of 12 years much later than in adults and adolescents, and the emergence of recent variants reduced vaccine effectiveness. Note also that the dosages for children under 12 years of age were lower than those of older individuals.

Our observation that bivalent boosters were more effective than monovalent boosters in children 5–11 years of age was consistent with findings from a recent study in adults and adolescents.^13^ However, bivalent boosters were not authorized in children 5–11 years of age until October 12, 2022. Thus, the number of children receiving bivalent boosters was small, and the follow-up was short.

Immunity acquired via omicron infection against omicron reinfection appeared similar between children under the age of 12 years and older individuals.^8^ However, the effectiveness of omicron infection against reinfection in adults and adolescents was studied over a relatively short time period.^8^

Vaccination rates were much lower in children under 12 years of age than in adults and adolescents, such that there was a great potential for selection bias in our study. Immuno-compromised children were more likely to be vaccinated than immuno-competent children, which would dilute vaccine effectiveness. Indeed, the third dose was part of the primary series for immune-compromised children rather than a booster dose. We did not have the information to separate additional doses for immune-compromised children from booster doses, such that our estimates of booster effectiveness pertained to third doses, which might not be booster doses.

Lastly, at-home SARS-CoV-2 tests became more common over the course of this study. Our database did not include home-based testing results and thus under-represented the cases of SARS-CoV-2 infection. In addition, hospitalization and survival status was not ascertained for all infected children. Our estimates of vaccine effectiveness would be biased if there were differential ascertainments of Covid-19 cases between vaccinated and unvaccinated children.

## Data Availability

All data produced in the present work are contained in the manuscript

## Supplementary Appendix

**Table S1.**
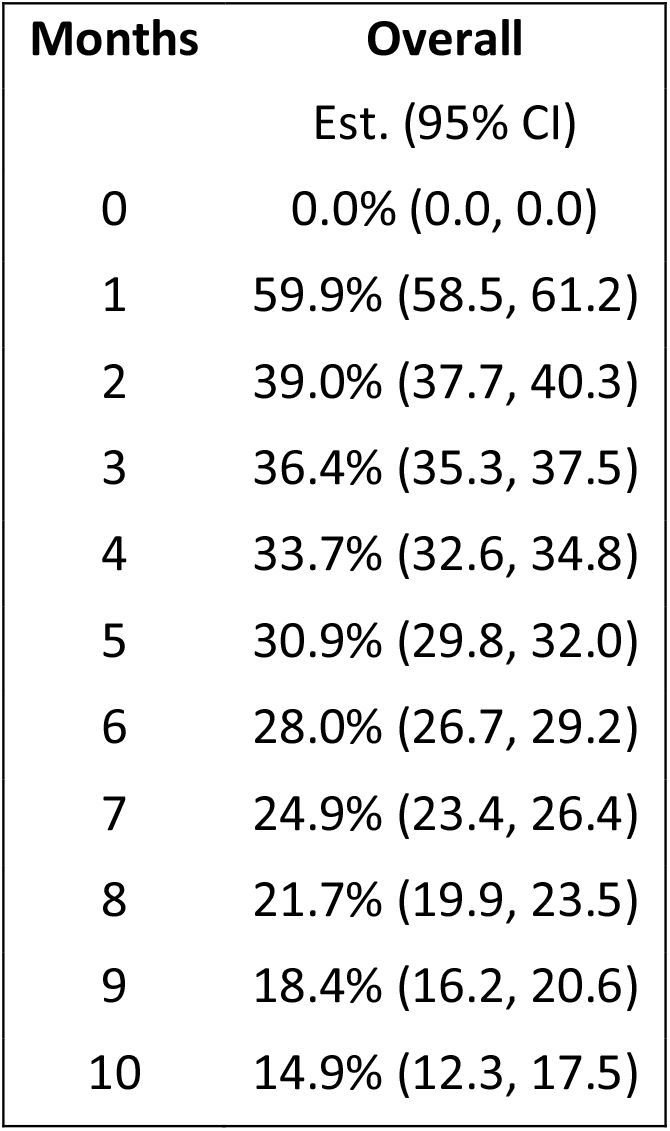
Estimates (95% CI) for the Effectiveness of Two Doses of an mRNA Vaccine, Compared with No Vaccination, against Infection, as a Function of Time Since First Dose, in Children 5 to 11 Years of Age.

| **Months** | **Overall**<br>**Est. (95% CI)** |
| --- | --- |
| 0 | 0.0% (0.0, 0.0) |
| 1 | 59.9% (58.5, 61.2) |
| 2 | 39.0% (37.7, 40.3) |
| 3 | 36.4% (35.3, 37.5) |
| 4 | 33.7% (32.6, 34.8) |
| 5 | 30.9% (29.8, 32.0) |
| 6 | 28.0% (26.7, 29.2) |
| 7 | 24.9% (23.4, 26.4) |
| 8 | 21.7% (19.9, 23.5) |
| 9 | 18.4% (16.2, 20.6) |
| 10 | 14.9% (12.3, 17.5) |

**Table S2.** Estimates (95% CI) for the Effectiveness of Two Doses of an mRNA Vaccine, Compared with No Vaccination, against Infection, as a Function of Time Since First Dose, by Date of First Dose, in Children 5 to 11 Years of Age.

| Months | Nov 2021 | Dec 2021 | Jan 2022 | Feb–May 2022 | June 2022–Jan 2023 |
| --- | --- | --- | --- | --- | --- |
|  | Est. (95% CI) | Est. (95% CI) | Est. (95% CI) | Est. (95% CI) | Est. (95% CI) |
| 0 | 0.0% (0.0, 0.0) | 0.0% (0.0, 0.0) | 0.0% (0.0, 0.0) | 0.0% (0.0, 0.0) | 0.0% (0.0, 0.0) |
| 1 | 77.3% (75.4, 79.0) | 59.4% (57.2, 61.5) | 66.9% (62.9, 70.5) | 58.6% (41.3, 70.8) | 68.9% (57.3, 77.3) |
| 2 | 35.0% (33.5, 36.4) | 38.6% (34.3, 42.7) | 42.8% (35.4, 49.3) | 55.4% (49.0, 61.1) | 56.4% (45.4, 65.1) |
| 3 | 32.6% (31.3, 33.9) | 36.7% (32.9, 40.2) | 40.1% (33.8, 45.9) | 52.0% (46.5, 56.9) | 48.9% (39.7, 56.6) |
| 4 | 30.1% (28.8, 31.4) | 34.6% (31.3, 37.7) | 37.3% (32.0, 42.2) | 48.3% (43.6, 52.6) | 40.1% (30.6, 48.2) |
| 5 | 27.6% (26.2, 28.9) | 32.5% (29.5, 35.3) | 34.4% (29.9, 38.6) | 44.3% (40.0, 48.2) | 29.7% (15.7, 41.5) |
| 6 | 24.9% (23.4, 26.4) | 30.3% (27.5, 33.0) | 31.3% (27.4, 35.1) | 39.9% (35.3, 44.3) |  |
| 7 | 22.1% (20.3, 23.9) | 28.0% (25.0, 30.9) | 28.1% (24.0, 32.1) | 35.3% (29.3, 40.8) |  |
| 8 | 19.3% (17.1, 21.4) | 25.7% (22.2, 29.1) | 24.8% (19.7, 29.6) | 30.3% (22.1, 37.6) |  |
| 9 | 16.3% (13.6, 18.8) | 23.3% (19.0, 27.3) | 21.3% (14.6, 27.4) | 24.9% (13.8, 34.6) |  |
| 10 | 13.2% (10.0, 16.2) | 20.8% (15.6, 25.7) | 17.6% (8.9, 25.5) | 19.1% (4.3, 31.5) |  |
| 11 | 10.0% (6.3, 13.6) | 18.2% (11.9, 24.2) | 13.7% (2.6, 23.6) |  |  |
| 12 | 6.7% (2.3, 10.9) | 15.6% (8.0, 22.6) |  |  |  |
| 13 | 3.2% (-1.8, 8.1) |  |  |  |  |

**Table S3.** Estimates (95% CI) for the Effectiveness of Two Doses of an mRNA Vaccine, Compared with No Vaccination, against Infection, as a Function of Time Since First Dose, by Previous Infection Status, in Children 5 to 11 Years of Age.

| Months | Uninfected<br>Est. (95% CI) | Infected<br>Est. (95% CI) |
| --- | --- | --- |
| 0 | 0.0% (0.0, 0.0) | 0.0% (0.0, 0.0) |
| 1 | 59.7% (58.3, 61.0) | 65.3% (58.1, 71.2) |
| 2 | 39.3% (38.0, 40.6) | 31.0% (23.1, 38.1) |
| 3 | 36.8% (35.7, 38.0) | 26.4% (19.2, 33.0) |
| 4 | 34.2% (33.1, 35.3) | 21.5% (14.9, 27.6) |
| 5 | 31.5% (30.3, 32.6) | 16.2% (10.0, 22.0) |
| 6 | 28.6% (27.3, 29.9) | 10.6% (4.4, 16.4) |
| 7 | 25.6% (24.0, 27.2) | 4.6% (-2.1, 10.9) |
| 8 | 22.5% (20.6, 24.4) |  |
| 9 | 19.3% (16.9, 21.6) |  |
| 10 | 16.0% (13.1, 18.7) |  |

**Table S4.** Estimates (95% CI) for the Effectiveness of Two Doses of an mRNA Vaccine, Compared with No Vaccination, against Hospitalization and Death, as a Function of Time Since First Dose, in Children 5 to 11 Years of Age.

| Months | Overall |
| --- | --- |
|  | Est. (95% CI) |
| 0 | 0.0% (0.0, 0.0) |
| 1 | 73.3% (8.3, 92.3) |
| 2 | 48.1% (-5.5, 74.5) |
| 3 | 39.3% (-11.5, 67.0) |
| 4 | 29.0% (-19.8, 57.9) |
| 5 | 17.0% (-32.0, 47.8) |
| 6 | 2.9% (-50.8, 37.4) |

**Table S5.**
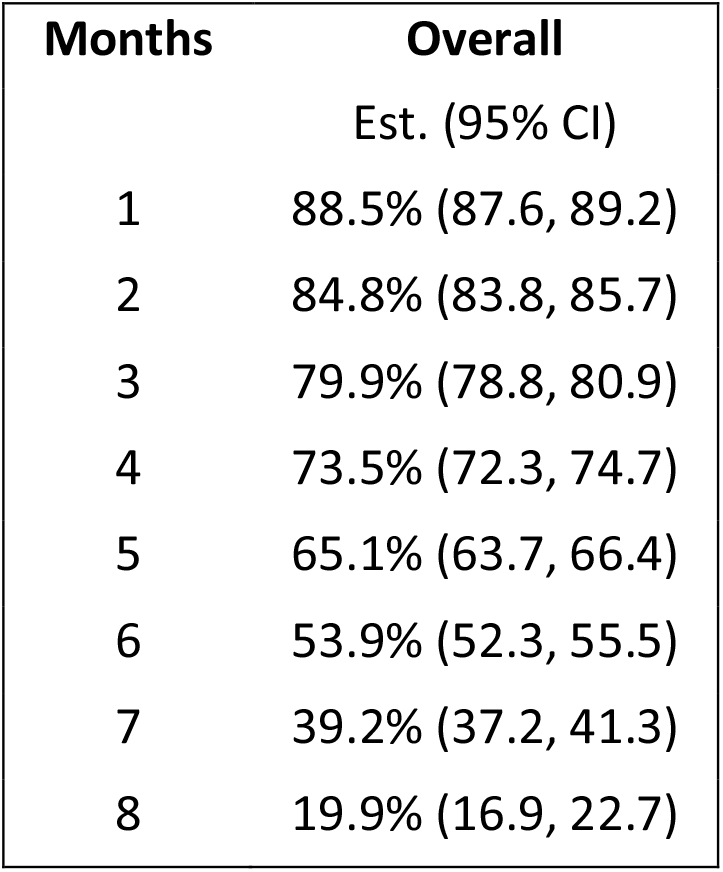
Estimates (95% CI) for the Effectiveness of Previous Omicron Infection, Compared with No Previous Infection, against Omicron Reinfection, as a Function of Time Since Previous Infection, in Children 5 to 11 Years of Age.

**Table S6.**
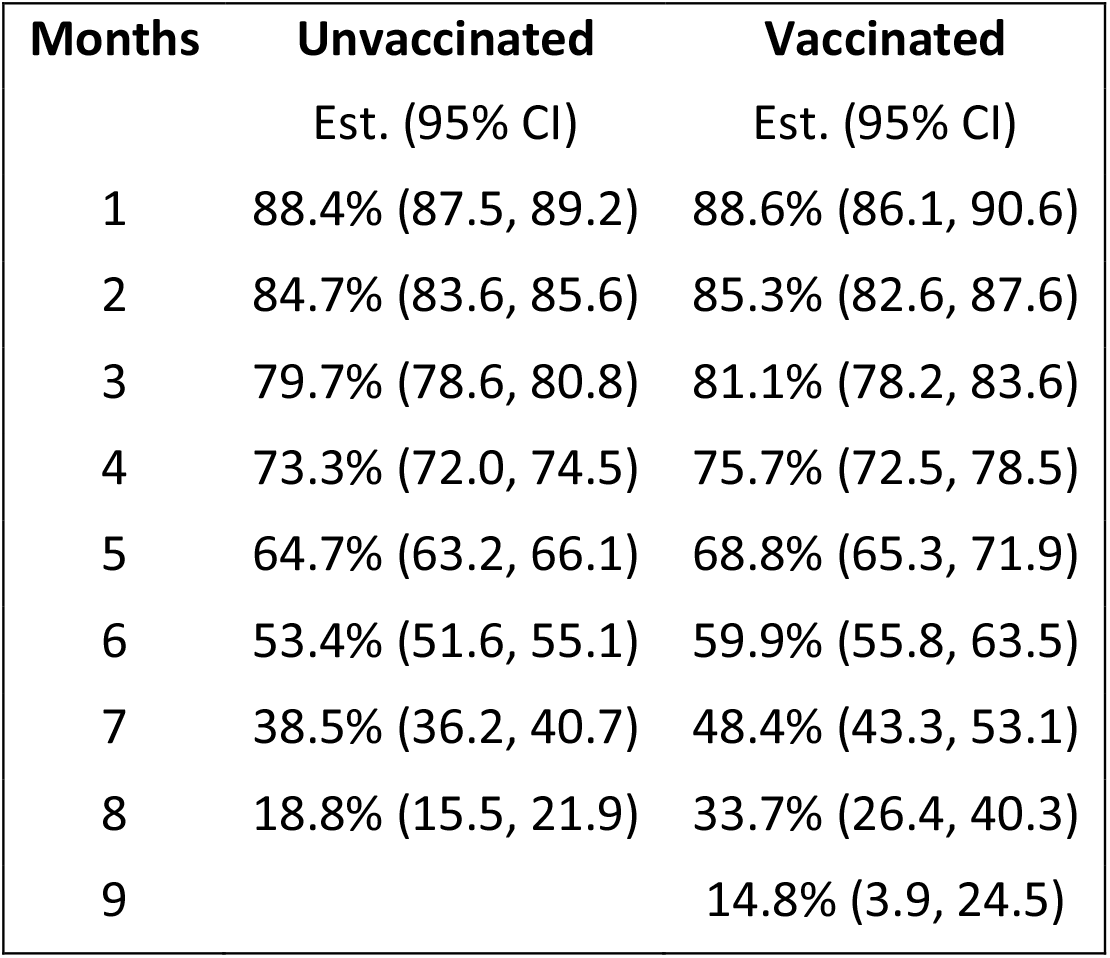
Estimates (95% CI) for the Effectiveness of Previous Omicron Infection, Compared with No Previous Infection, against Omicron Reinfection, as a Function of Time Since Previous Infection, by Vaccination Status, in Children 5 to 11 Years of Age.

| **Months** | **Unvaccinated** | **Vaccinated** |
| --- | --- | --- |
|  | Est. (95% CI) | Est. (95% CI) |
| 1 | 88.4% (87.5, 89.2) | 88.6% (86.1, 90.6) |
| 2 | 84.7% (83.6, 85.6) | 85.3% (82.6, 87.6) |
| 3 | 79.7% (78.6, 80.8) | 81.1% (78.2, 83.6) |
| 4 | 73.3% (72.0, 74.5) | 75.7% (72.5, 78.5) |
| 5 | 64.7% (63.2, 66.1) | 68.8% (65.3, 71.9) |
| 6 | 53.4% (51.6, 55.1) | 59.9% (55.8, 63.5) |
| 7 | 38.5% (36.2, 40.7) | 48.4% (43.3, 53.1) |
| 8 | 18.8% (15.5, 21.9) | 33.7% (26.4, 40.3) |
| 9 |  | 14.8% (3.9, 24.5) |

**Table S7.** Estimates (95% CI) for the Effectiveness of Previous Infection with the Delta Variant, Compared with No Previous Infection, against Reinfection, as a Function of Time Since Previous Infection, by Vaccination Status, in Children 5 to 11 Years of Age.

| **Months** | **Unvaccinated** | **Vaccinated** |
| --- | --- | --- |
|  | Est. (95% CI) | Est. (95% CI) |
| 1 | 84.7% (83.9, 85.5) | 88.8% (78.6, 94.2) |
| 2 | 82.5% (81.6, 83.3) | 85.9% (75.0, 92.1) |
| 3 | 79.9% (79.1, 80.7) | 82.3% (70.5, 89.4) |
| 4 | 77.0% (76.1, 77.8) | 77.7% (65.0, 85.8) |
| 5 | 73.7% (72.8, 74.5) | 71.9% (57.9, 81.2) |
| 6 | 69.8% (68.9, 70.8) | 64.6% (48.4, 75.7) |
| 7 | 65.5% (64.4, 66.5) | 55.4% (35.3, 69.2) |
| 8 | 60.5% (59.2, 61.7) | 43.7% (17.0, 61.9) |
| 9 | 54.7% (53.1, 56.2) | 29.1% (-8.7, 53.8) |
| 10 | 48.1% (46.1, 50.1) | 10.7% (-44.5, 44.8) |

**Table S8.** Estimates (95% CI) for the Effectiveness of Previous Infection, Compared with No Previous Infection, against Reinfection Resulting in Hospitalization or Death, as a Function of Time Since Previous Infection, in Children 5 to 11 Years of Age.

| **Months** | **Overall** |
| --- | --- |
|  | Est. (95% CI) |
| 1 | 87.5% (64.2, 95.6) |
| 2 | 85.7% (61.8, 94.7) |
| 3 | 83.8% (59.1, 93.6) |
| 4 | 81.6% (56.2, 92.2) |
| 5 | 79.0% (53.0, 90.6) |
| 6 | 76.2% (49.4, 88.8) |
| 7 | 72.9% (45.2, 86.6) |
| 8 | 69.2% (40.5, 84.0) |
| 9 | 64.9% (34.9, 81.1) |
| 10 | 60.1% (28.4, 77.8) |

**Table S9.** Estimates (95% CI) for the Effectiveness of a Monovalent or a Bivalent Booster Dose, Compared with Two Doses, against Omicron Infection, as a Function of Time since Receipt of Booster Dose, in Children 5 to 11 Years of Age.

| **Months** | **Monovalent** | **Bivalent** |
| --- | --- | --- |
|  | Est. (95% CI) | Est. (95% CI) |
| 0 | 0.0% (0.0, 0.0) | 0.0% (0.0, 0.0) |
| 1 | 24.4% (14.4, 33.2) | 76.7% (45.7, 90.0) |
| 2 | 23.1% (15.6, 30.0) | 47.3% (-17.9, 76.4) |
| 3 | 21.9% (14.9, 28.3) |  |
| 4 | 20.6% (11.7, 28.6) |  |
| 5 | 19.3% (6.9, 30.0) |  |

**Table S10.** Estimates (95% CI) for the Effectiveness of Two Doses of an mRNA Vaccine, Compared with No Vaccination, against Infection, as a Function of Time Since First Dose, in Children 0 to 4 Years of Age.

| Months | Overall |
| --- | --- |
|  | Est. (95% CI) |
| 0 | 0.0% (0.0, 0.0) |
| 1 | 55.6% (49.2, 61.2) |
| 2 | 63.8% (57.0, 69.5) |
| 3 | 62.0% (57.4, 66.0) |
| 4 | 60.1% (54.4, 65.1) |
| 5 | 58.1% (48.3, 66.1) |

**Table S11.** Estimates (95% CI) for the Effectiveness of Two Doses of an mRNA Vaccine, Compared with No Vaccination, against Infection, as a Function of Time Since First Dose, by Date of First Dose, in Children 0 to 4 Years of Age.

| **Months** | **June 2022** | **July 2022** | **Aug 2022–Jan 2023** |
| --- | --- | --- | --- |
|  | Est. (95% CI) | Est. (95% CI) | Est. (95% CI) |
| 0 | 0.0% (0.0, 0.0) | 0.0% (0.0, 0.0) | 0.0% (0.0, 0.0) |
| 1 | 56.7% (43.6, 66.8) | 59.1% (47.5, 68.1) | 59.9% (48.9, 68.6) |
| 2 | 62.7% (51.5, 71.3) | 69.6% (59.5, 77.2) | 58.8% (51.4, 65.0) |
| 3 | 61.9% (53.9, 68.6) | 67.1% (59.8, 73.0) | 57.6% (49.3, 64.5) |
| 4 | 61.2% (54.1, 67.2) | 64.3% (57.8, 69.8) | 56.4% (42.7, 66.7) |
| 5 | 60.4% (51.5, 67.7) | 61.3% (52.0, 68.9) |  |
| 6 | 59.7% (46.6, 69.5) |  |  |

**Table S12.** Estimates (95% CI) for the Effectiveness of Two Doses of an mRNA Vaccine, Compared with No Vaccination, against Infection, as a Function of Time Since First Dose, by Vaccine Product, in Children 0 to 4 Years of Age.

| **Months** | **BNT162b2** | **mRNA-1273** |
| --- | --- | --- |
|  | Est. (95% CI) | Est. (95% CI) |
| 0 | 0.0% (0.0, 0.0) | 0.0% (0.0, 0.0) |
| 1 | 54.2% (45.8, 61.2) | 58.0% (47.5, 66.5) |
| 2 | 63.3% (54.3, 70.5) | 64.4% (53.2, 73.0) |
| 3 | 63.5% (57.8, 68.4) | 59.5% (51.6, 66.1) |
| 4 | 63.7% (56.7, 69.5) | 53.9% (43.6, 62.4) |
| 5 | 63.9% (52.2, 72.7) | 47.6% (27.7, 62.0) |

**Table S13.**
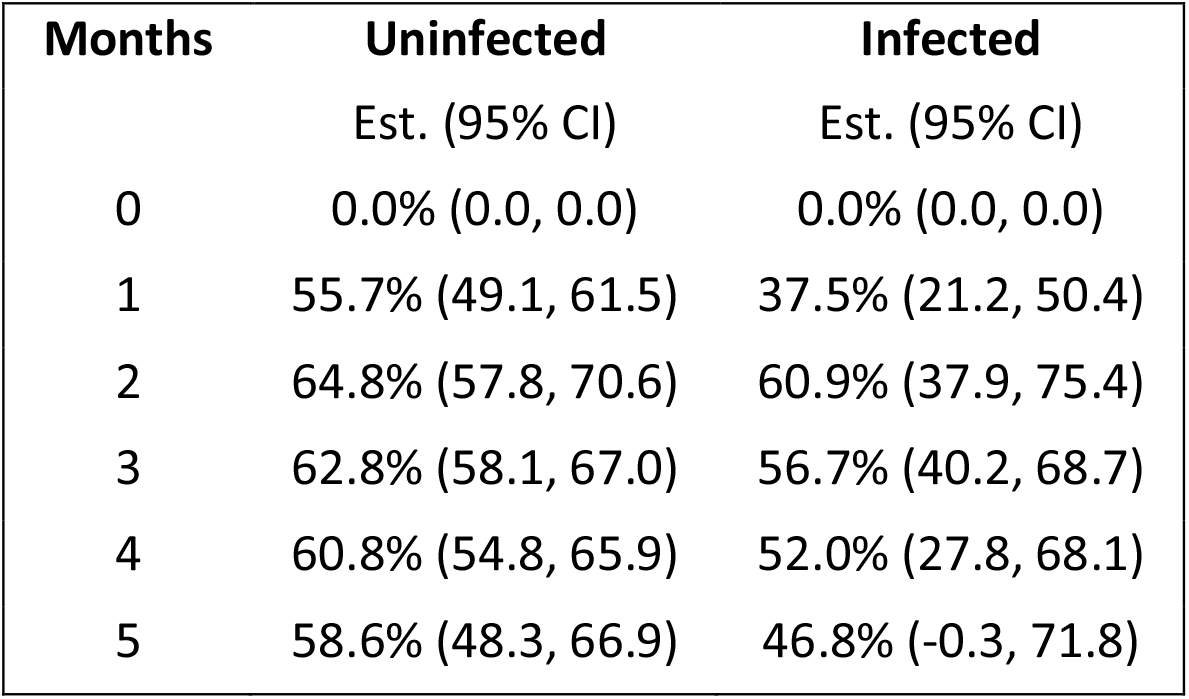
Estimates (95% CI) for the Effectiveness of Two Doses of an mRNA Vaccine, Compared with No Vaccination, against Infection, as a Function of Time Since First Dose, by Previous Infection Status, in Children 0 to 4 Years of Age.

**Table S14.** Estimates (95% CI) for the Effectiveness of Previous Omicron Infection, Compared with No Previous Infection, against Reinfection, as a Function of Time Since Previous Infection, in Children 0 to 4 Years of Age.

| **Months** | **Overall** |
| --- | --- |
|  | Est. (95% CI) |
| 1 | 83.1% (81.7, 84.3) |
| 2 | 80.4% (79.0, 81.7) |
| 3 | 77.3% (75.9, 78.6) |
| 4 | 73.7% (72.4, 75.0) |
| 5 | 69.6% (68.2, 70.9) |
| 6 | 64.7% (63.3, 66.1) |
| 7 | 59.2% (57.5, 60.8) |
| 8 | 52.7% (50.6, 54.8) |
| 9 | 45.2% (42.4, 48.0) |
| 10 | 36.6% (32.7, 40.2) |

**Table S15.** Estimates (95% CI) for the Effectiveness of Previous Infection, Compared with No Previous Infection, against Reinfection Resulting in Hospitalization or Death, as a Function of Time Since Previous Infection, in Children 0 to 4 Years of Age.

| **Months** | **Overall** |
| --- | --- |
|  | Est. (95% CI) |
| 1 | 63.3% (26.1, 81.8) |
| 2 | 62.4% (27.8, 80.4) |
| 3 | 61.4% (29.4, 78.9) |
| 4 | 60.4% (30.6, 77.4) |
| 5 | 59.4% (31.6, 75.9) |
| 6 | 58.4% (32.2, 74.5) |
| 7 | 57.3% (32.4, 73.1) |
| 8 | 56.2% (32.0, 71.8) |
| 9 | 55.1% (31.1, 70.8) |
| 10 | 54.0% (29.5, 70.0) |

